# The impact of Long COVID on Health-Related Quality-of-Life using OpenPROMPT

**DOI:** 10.1101/2023.12.06.23299601

**Authors:** Oliver Carlile, Andrew Briggs, Alasdair D Henderson, Ben FC Butler-Cole, John Tazare, Laurie A Tomlinson, Michael Marks, Mark Jit, Liang-Yu Lin, Chris Bates, John Parry, Sebastian CJ Bacon, Iain Dillingham, William A Dennison, Ruth E Costello, Alex J Walker, William Hulme, Ben Goldacre, Amir Mehrkar, Brian MacKenna, The OpenSAFELY Collaborative, Emily Herrett, Rosalind M Eggo

**Affiliations:** London School of Hygiene and Tropical Medicine, Keppel Street, London WC1E 7HT, UK; Bennett Institute for Applied Data Science, Nuffield Department of Primary Care Health Sciences, University of Oxford, OX2 6GG, UK; TPP, TPP House, 129 Low Lane, Horsforth, Leeds, LS18 5PX, UK; Patient and Public Involvement Steering Committee, London, UK; Hospital for Tropical Diseases, University College London Hospital, London, WC1E 6JD, UK; Division of Infection and Immunity, University College London, London, WC1E 6BT, UK

## Abstract

**Background:** Long COVID is a major problem affecting patient health, the health service, and the workforce. To optimise the design of future interventions against COVID-19, and to better plan and allocate health resources, it is critical to quantify the health and economic burden of this novel condition.

**Methods:** With the approval of NHS England, we developed OpenPROMPT, a UK cohort study measuring the impact of long COVID on health-related quality-of-life (HRQoL). OpenPROMPT invited responses to Patient Reported Outcome Measures (PROMs) using a smartphone application and recruited between November 2022 and October 2023. We used the validated EuroQol EQ-5D questionnaire with the UK Value Set to develop disutility scores (1-utility) for respondents with and without Long COVID using linear mixed models, and we calculated subsequent Quality-Adjusted Life-Months (QALMs) for long COVID.

**Results:** We used data from 6,070 participants where 24.7% self-reported long COVID. In multivariable regressions, long COVID had a consistent impact on HRQoL, showing a high probability of reporting loss in quality-of-life (OR: 22, 95% CI:12.35-39.29) compared with people who did not report long COVID. Reporting a disability was the largest predictor of losses of HRQoL (OR: 60.2, 95% CI: 27.79-130.57) across survey responses. Self-reported long COVID was associated with an 0.37 QALM loss.

**Conclusions:** We found substantial impacts on quality-of-life due to long COVID, representing a major burden on patients and the health service. We highlight the need for continued support and research for long COVID, as HRQoL scores compared unfavourably to patients with conditions such as multiple sclerosis, heart failure, and renal disease.

## Introduction

Following infection by SARS-CoV-2, the majority of patients will recover within 4 weeks but 10-15% do not, and may face significant impacts on their health-related quality-of-life (HRQoL) (Davis et al., 2023). The National Institute for Health and Care Excellence (NICE) in the UK developed three clinical definitions for the effects following infection: ‘acute COVID-19’ for signs and symptoms of COVID-19 between 0 and 4 weeks, ‘ongoing symptomatic COVID-19’ for symptoms between 4 to 12 weeks, and ‘post COVID-19 syndrome’ with symptoms persisting 12 weeks or longer not explained by an alternative diagnosis (NICE, 2020). The latter two definitions refer to long COVID.

Persistent symptoms reported to occur after infection are wide-ranging, with the most common being fatigue, shortness of breath, muscular, joint and chest pains, headaches, persistent cough, and altered senses of smell and taste (Aiyegbusi et al., 2021). As of 2nd January 2023, the Office for National Statistics (2023) figures for the prevalence of self-reported long COVID estimated 2 million people in the UK were experiencing symptoms persisting longer than four weeks, not explained by other diagnoses. An estimated 1.5 million people (77%) with self-reported long COVID reported symptoms adversely affected day-to-day activities, and 380,000 (19%) reported ability to undertake day-to-day activities had been ‘limited a lot’ (ONS, 2023). With an estimated 22.2 million UK cases of COVID-19 as of May 2022, the burden of long COVID may be wide ranging for the NHS (GOV.UK, 2023). With the extensive symptoms of long COVID, the impact on HRQoL can be significant, with Walker et al. (2023) estimating EQ-5D scores among patients referred to post-COVID clinics in England and Wales were worse than those among patients with metastatic cancers.

Few studies have assessed quality adjusted life years (QALYs) attributable to long COVID as most focused on the effect of acute COVID-19 on HRQoL. Sigfrid et al., (2021) examined EQ-5D-5L survey results in the UK at least 90 days after suspected SARS-CoV-2 hospitalisation between 17th January to 5th October 2020. 54% reported they had not fully recovered at time of follow-up, with 93% reporting persistent symptoms. Previous studies of QALYs lost due to COVID were limited to small numbers of respondents, for example in Sandmann et al. (2022) estimating losses for 548 positive cases against a control group of 651 respondents, and have not explored inequalities by patient characteristics.

This study addresses this gap, specifically identifying the impact of long COVID, the contribution of symptom-specific patient-reported outcomes measures (PROMs) to assessment of quality-of-life, and aims to quantify how this results in QALY losses.

## Methods

### OpenPROMPT

We conducted a cohort study using *Airmid*, the in-house smartphone application of TPP, which is the software provider for 40% of all primary care providers in England (Andrews et al., 2022). Full details of the study protocol and methods have been previously published (Herrett et al., 2023). In brief, any adult in England was eligible to participate in the study, if they were able to download and use the *Airmid* app and consented to the study (Herrett et al., 2023). Participants were requested to fill questionnaires in 30-day intervals: day 0 (the point of enrolment in the study), then days 30, 60, and 90. Survey responses were categorised as falling in these points of time if completed +/-5 days of the 30-day intervals. There was also a questionnaire at recruitment collecting demographic information. Recruitment took place between November 11^th^ 2022 and July 31^st^ 2023.

The questionnaires consisted of existing validated PROMS which covered a range of themes, with the EuroQol EQ-5D-5L the primary outcome measure for this study. To assess the impact of long COVID on other aspects of HRQoL, symptom specific questionnaires were used including the Medical Research Council (MRC) Dyspnoea breathlessness Scale (UKRI, 2016) and Functional Assessment of Chronic Illness Therapy – Fatigue (FACIT-F Fatigue) Scale (Tennant, 2011). Patient-reported responses to OpenPROMPT questionnaires were automatically linked to primary care records managed by TPP SystmOne if the patient was registered at a practice using TPP software, and were stored in the patient health record. We accessed these data via the OpenSAFELY research platform, where all data were linked, stored and analysed securely (https://opensafely.org/). All data, including coded diagnoses, medications and physiological parameters, are pseudonymised. No free text data were included.

Due to the difficulties in assessing history of long COVID from medical records, the experience of COVID-19 required a specific questionnaire (Walker et al., 2021). Patients were defined as self-reporting long COVID if they responded both “No I still have symptoms” to the question “Thinking of your last episode of COVID-19, have you now recovered to normal?” and secondly that symptoms lasted either 4-12 weeks, or more than 12 weeks to the question “How long have you had/did you have COVID-19 symptoms overall?”. Participants missing responses to both these questions were defined as not stated.

### Participant demographics and comorbidities

Participant characteristics were collected through the recruitment questionnaire and linked clinical records. OpenSAFELY contains electronic health records (EHRs) drawn from primary, secondary care (inpatient, outpatient, emergency) and all prescriptions, allowing in-depth assessment of patient comorbidities. The presence of pre-existing comorbidities at baseline was based upon previous research within OpenSAFELY on fifteen chronic comorbidities (Thompson et al., 2022). The Index of Multiple Deprivation (IMD) for participants was based on their postcode address at lower super output area. Specific assessments on the impact of COVID-19 were collected from EHRs, including diagnosis or referral codes for long COVID. The recruitment questionnaire collected ethnicity, education level, and annual household incomes. Age, in bands of 18-29, 30-39, 40-49, 50-59, 60-69, and 70+, sex and NHS region were extracted from the patients EHRs.

### Outcomes

The EQ-5D-5L is a standardised measure widely used to collect information on HRQoL across interventions and conditions (Devlin and Brooks, 2017). It asks respondents to describe their health on that day, covering five dimensions of quality-of-life: mobility, self-care, usual activities, pain and discomfort, and anxiety and depression. Each dimension has five possible responses: level 1: no problems, level 2: slight problems, level 3: moderate problems, level 4: severe problems, and level 5: extreme problems/unable to. Responses return a five-digit descriptive code for health state (e.g., 14523).

Secondary symptom specific PROMs were collected on Fatigue using the FACIT-F scale. Participants responded to 13 statements related to daily functioning and activities with a 7 day recall period.

We assessed breathlessness using the MRC Dyspnoea Scale, which records the degree of breathlessness relating to daily activities with no recall period. The scale defines grade 1 as mild, grades 2-3 as moderate, and grades 4-5 as severe breathlessness.

### EuroQol EQ-5D Score

To estimate EQ-5D score, we used the EuroQol mapping function defined within Hernández Alava et al, (2023) to obtain utility values from the three level (3L) UK value set by mapping to the five level (5L) format collected in OpenPROMPT using a development of the van Hout (2012) crosswalk function. The value set for the UK is derived from population-based studies of valuation for health states using time-trade-off, resulting in a preference based score for each health state. Perfect health with no problems in any dimensions of quality-of-life (i.e., 11111) returns a score of 1, with death anchored at zero and states deemed worse than death returning negative scores. We used the utility score to generate disutility (1-EQ5D utility) as the lost quality-of-life from a perfect health state. Only data which was linked to TPP medical records were used, as the mapping function requires age to derive utility scores. We excluded participants from analysis who self-defined as non-binary gender because the function accounts for only male/female responses.

## Statistical Analysis

### HRQoL EQ-5D Disutility Score

We used multivariable regression models for the impact of long COVID on loss of utility from perfect health, referred to as disutility and measured as 1 minus the EQ-5D-5L utility value. To handle individuals with no alteration to HRQoL, we used a two-part model, first modelling the probability of any impact on quality-of-life and secondly the effect on quality-of-life. The first part of the model was a mixed effect logistic regression on the probability of returning disutility greater than zero, indicating loss of HRQoL, with adjustment for within-participant correlation of EQ-5D-5L responses across surveys. The second part of the model employed mixed effects linear models on disutility. Missing data, from non-response or loss to follow-up were assumed to be missing at random, where linear mixed models are appropriate alternatives to imputation when missingness is high (Gabrio et al., 2022). Models were adjusted for demographic indicators including age, sex, ethnicity and IMD quintiles, and used to assess the impact of variables such as household income, education and previous COVID hospitalisations on the outcome by inclusion in the mixed models.

Subsequent models included the secondary PROMs (MRC-Dyspnoea Scale and FACIT-F scores) to explore their contribution to long COVID related losses in HRQoL. To match the relationship with disutility, we reversed the FACIT-F scores. With a maximum possible score of 52, higher values now indicate greater fatigue. We sequentially added the PROMs in a stepwise approach and assessed if overall model fit was improved based on the Akaike Information Criterion (AIC). This assessed the extent with which PROMs on symptom-specific issues influenced the overall HRQoL measured by EQ-5D-5L.

We compared characteristics of the cohort collected at baseline recruitment to those completing all surveys to assess the extent of selection bias attributable to missing EQ-5D-5L scores due to loss to follow-up.

### Quality-adjusted life-years (QALYs)

Using results from the longitudinal EQ-5D-5L survey responses, we extrapolated the responses to estimate the QALYs lost due to long COVID. QALYs were calculated using the area under the curve method at individual level using disutility scores (Hunter et al., 2015). The relationship between utility scores over time assumes linearity given the short duration between EQ-5D-5L measurements. Because respondents reported HRQoL for less than 12 months, we did not apply discounting to the total QALYs. The results are shown in quality-adjusted life-months (QALMs) which do not reshape the time aspect of QALYs in terms of years.

QALM losses were estimated using a complete case analysis with participants who completed all surveys, and we compared the results to available case analysis. We also separated by a long COVID diagnosis in the participant’s EHRs, with consistent evidence that individuals heavily impacted are more likely to respond in data collection (Sudre et al. 2021). We conducted linear regression models to assess associations between total QALMs lost, adjusting for age, sex, disability, the number of comorbidities and baseline utility.

Data management was performed using Python 3 in OpenSAFELY, with analysis conducted using Stata version 16.1. Code for data management and analysis, as well as codelists are online (<u>opensafely/openprompt-hrqol (github.com)</u>).

## Results

### Descriptive Analysis

Overall, 6070 participants with linked TPP EHRs completed the recruitment questionnaires and were included in the analysis. 61% were female, with a median age of 53 (IQR 43-62) and the majority of participants were White (5765/6045, 95%) (Table 1). 1495/3975 participants (24.7%) self-reported long COVID, but only 6% had a long COVID diagnosis in their EHR (Table 1). As not all questions were mandatory, we treated non-responses to both questions required for defining long COVID as not stated, corresponding to 2095/6070 (34.5%) participants.

**Table 1.**
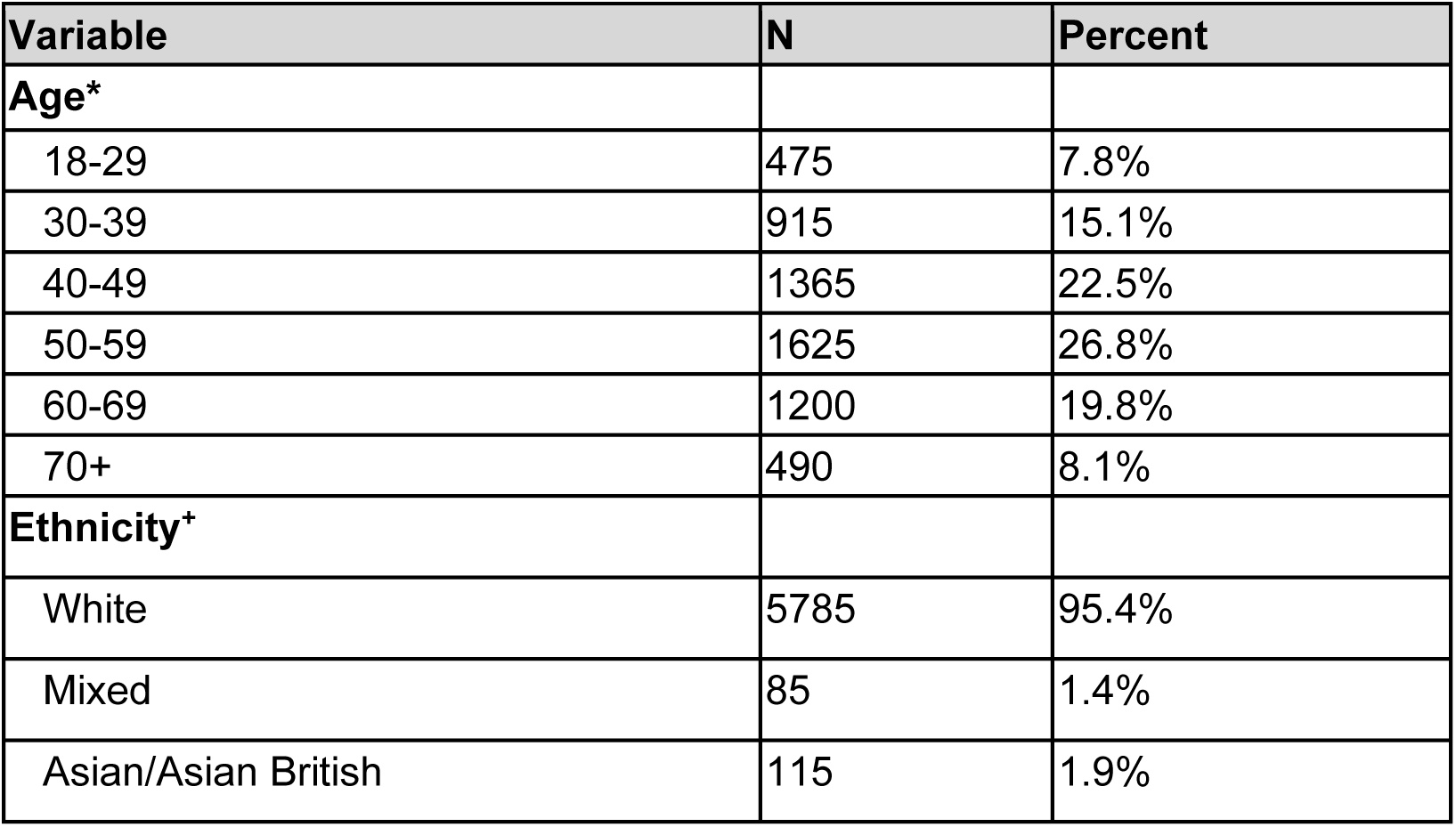
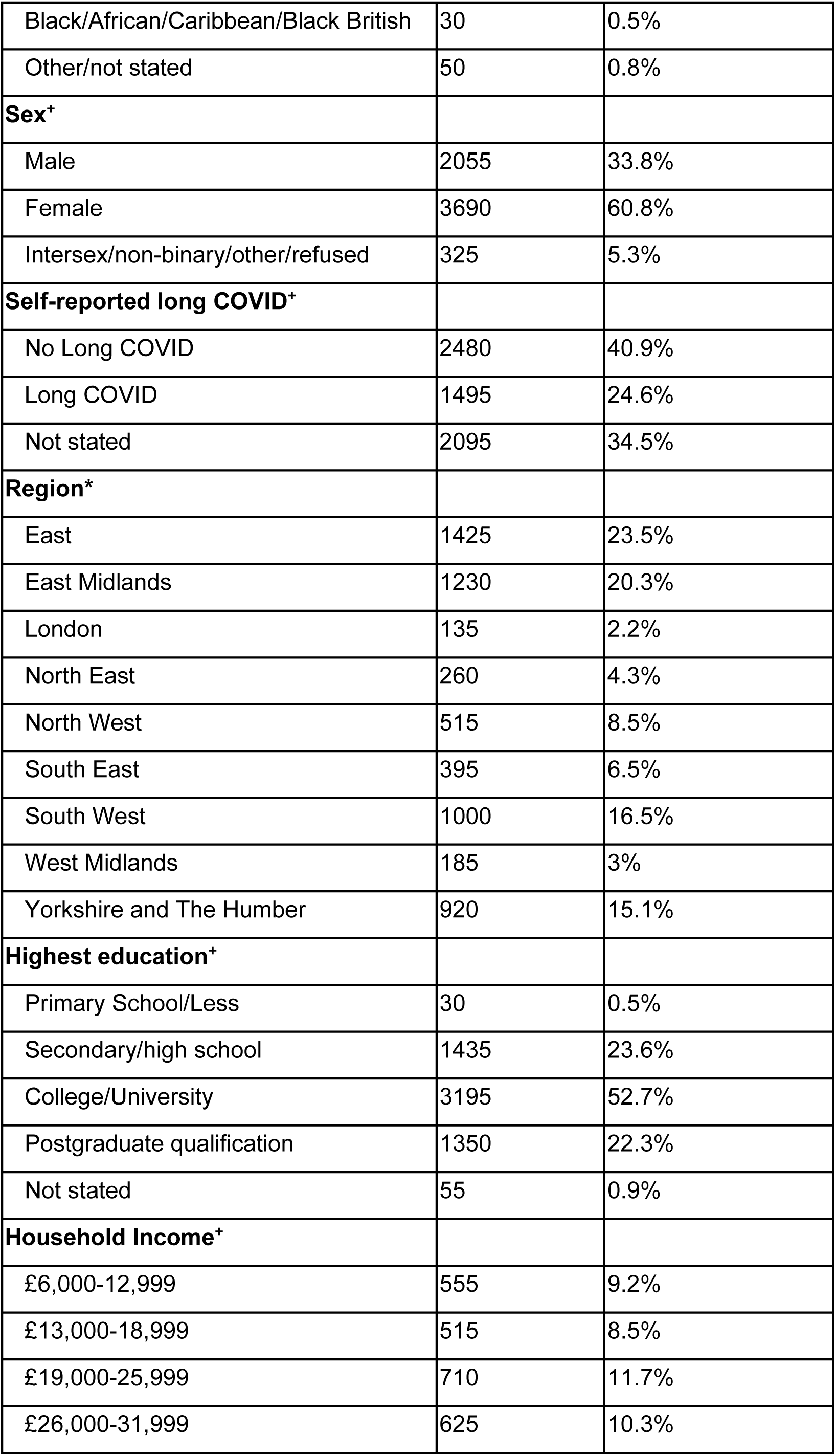
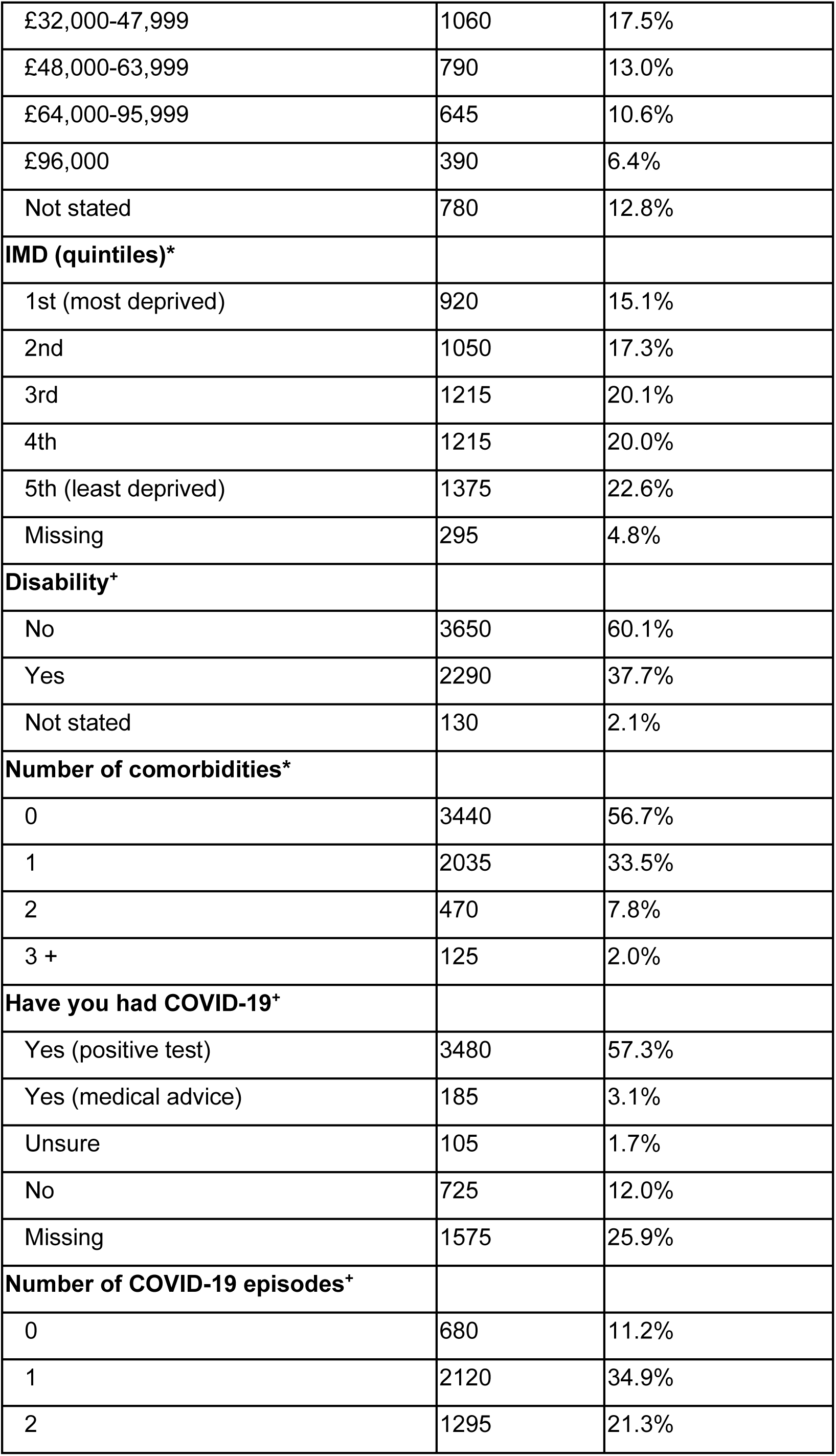
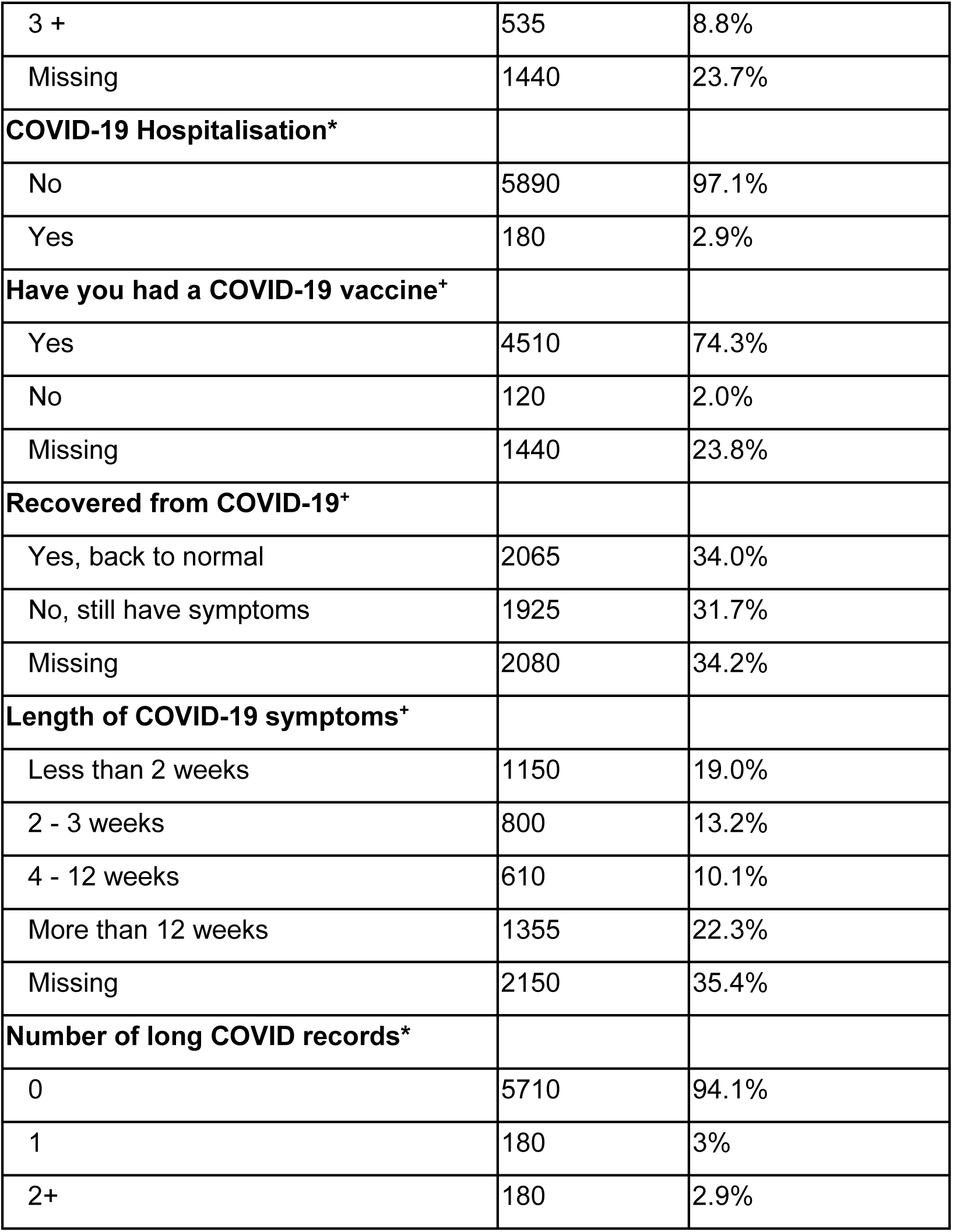
Demographic characteristics reported in recruitment surveys and EHRs. * indicates the use of EHRs and + indicates questionnaire responses.

705 respondents reported no problems across any dimensions of EQ-5D-5L at baseline. The distribution of EQ-5D disutility scores was positively skewed, with a small number (<50) having severe losses on HRQoL (Figure 1a). The distribution of participant responses to each dimension of EQ-5D-5L shows greater impact of long COVID for anxiety and depression and pain and discomfort, with little impact on mobility and self-care (Figure 1f).

**Figure 1.**
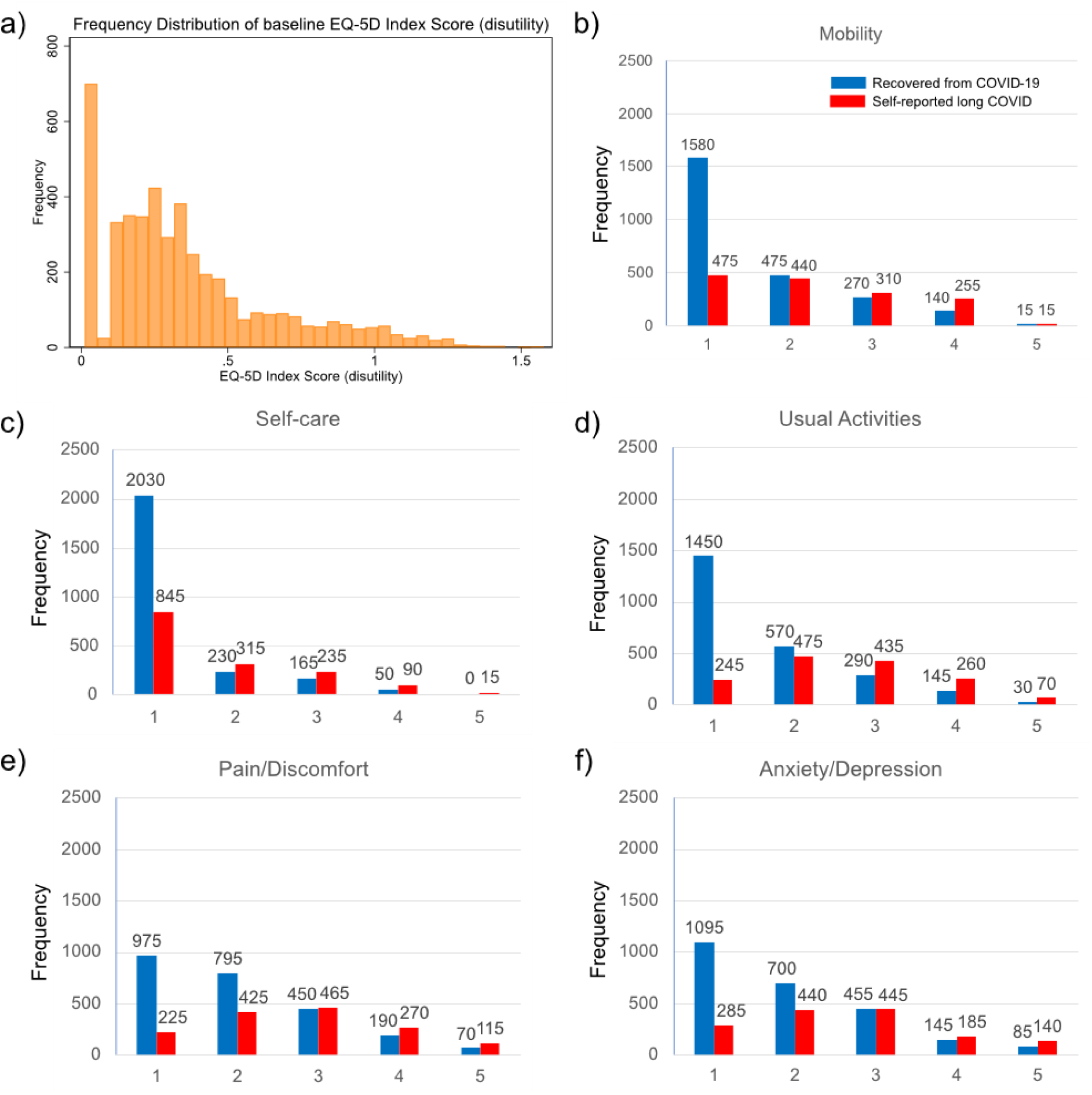
Self-reported quality of life measures. a) Frequency distribution of baseline EQ-5D-5L score (disutility), b) Mobility dimension of EQ-5D, c) Self-care dimension of EQ-5D, d) Usual activities dimension of EQ-5D. e) Pain/discomfort dimension of EQ-5D, f) Anxiety/depression dimension of EQ-5D. Each dimension has five possible responses: level 1: no problems, level 2: slight problems, level 3: moderate problems, level 4: severe problems, and level 5: extreme problems/unable to. Blue marks the participant did not report long COVID, and red that they did. Responses are only shown for the 3975 non-missing self-reported long COVID respondents

### Health-Related Quality-of-Life

Participants self-reporting long COVID were highly likely to report loss of HRQoL compared to participants who did not report long COVID (OR 21.2 (12.03; 37.54) for returning a loss of HRQoL and 0.075 (0.06, 0.09) unit lower quality of life) (Figure 2). The largest odds ratio for reporting a loss of HRQoL was for disability, but there were also associations with presence of comorbidities and gender. Coefficients for HRQoL loss were higher in those with comorbidities and with lower incomes (Figure 2). Odds ratios for reporting any disutility and coefficients for distutility are given in Supplementary table 2.

**Figure 2.**
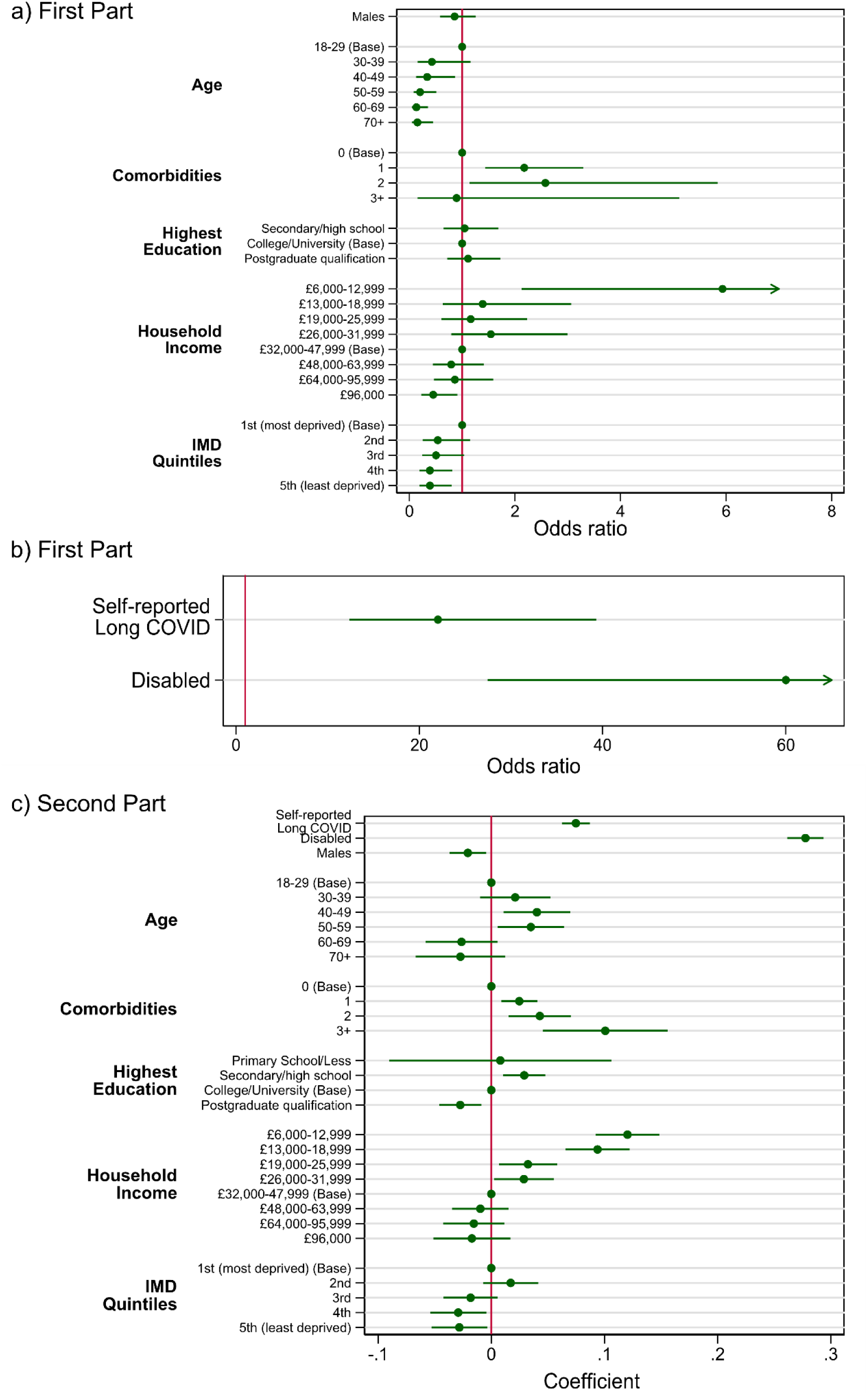
Model outputs for disutility. a) Odds ratios for the probability of reporting disutility in the first part of the full model. Note that greater odds ratio relates to a higher odds of reporting a negative change in HRQoL. b) Odds ratios for self-reported long COVID and disability in the first part of the model shown separately to allow visualisation, due to their much higher odds ratios. c) Coefficients for the second part of the model. Note that negative coefficients relate to lower disutility, i.e. higher quality-of-life.

**Figure 3.**
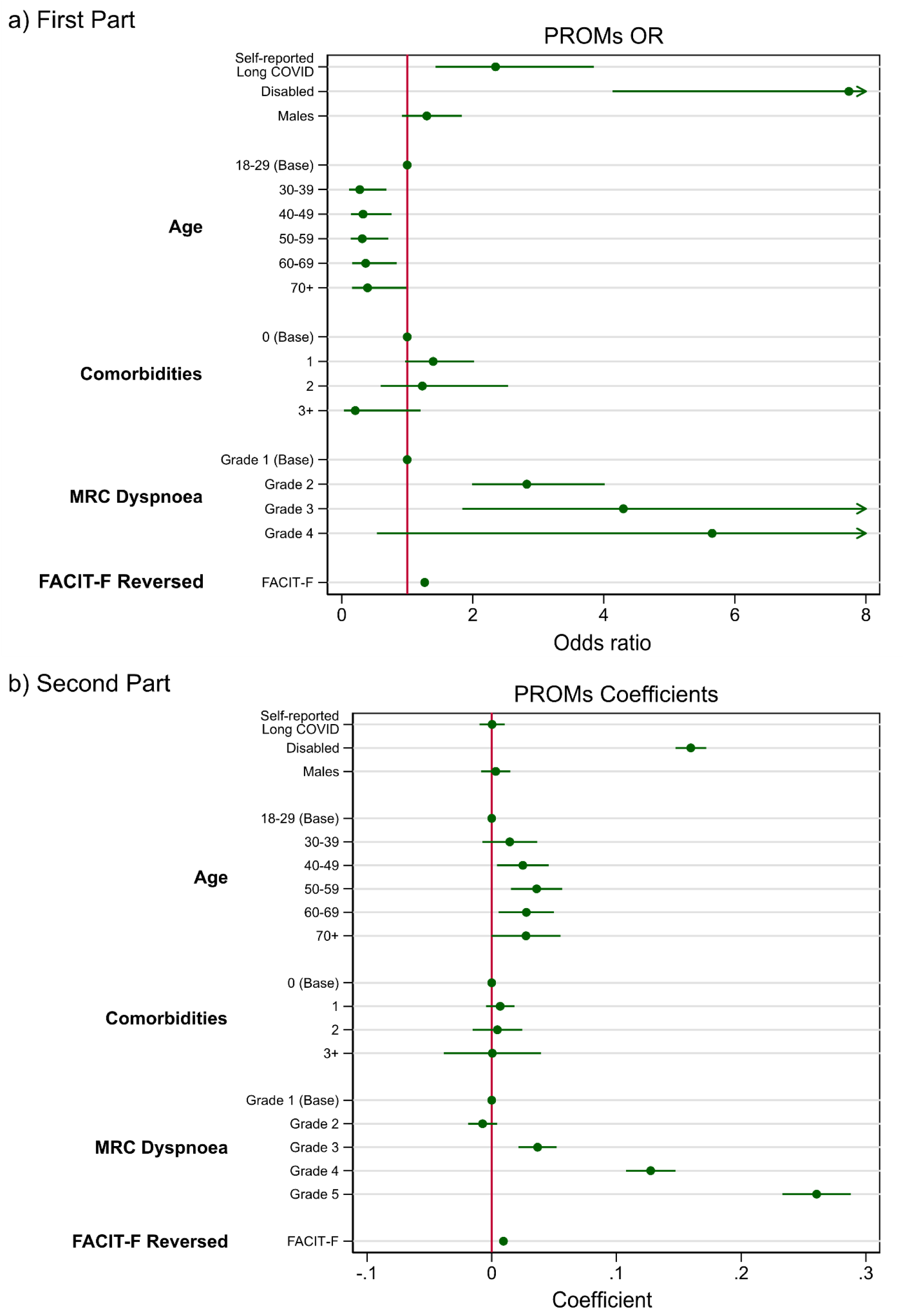
Model outputs for disutility including additional Patient-Reported Outcome Measures (PROMS). a) Odds ratios for the probability of reporting disutility in the first part of the full model including PROMs. Note that greater odds ratio relates to a higher odds of reporting a negative change in disutility. b) Coefficients for the second part of the model include PROMs. Note that negative coefficients relate to lower disutility, i.e. higher quality-of-life.

We found associations between the breathlessness and fatigue PROMs and HRQoL (Table 3). A unit increase in FACIT-F, i.e., reporting higher levels of fatigue, is estimated to have an odds ratio of 1.26 (95% CI:1.22; 1.31) for reporting loss in HRQoL. Severe grades of breathlessness (grades 4-5) resulted in an 0.13 (95% CI:0.11; 0.15) and 0.26 (95% CI:0.23; 0.29) loss of quality-of-life respectively, compared to reporting no effect, with the highest grade predicting HRQoL loss perfectly. After adjusting for breathlessness and fatigue, the remaining estimated reduction in HRQoL due to reported long COVID was lower. The OR fell from 22.0 (95% CI 12.4-39.2) to 2.35 (1.43-3.85), with the unit loss of HRQoL falling from 0.075 to 0.0003.

### Individual QALM Losses

People who self-reported long COVID had lower utility scores for every month after recruitment, with little change in HRQoL over time. (Figure 4a).

**Figure 4.**
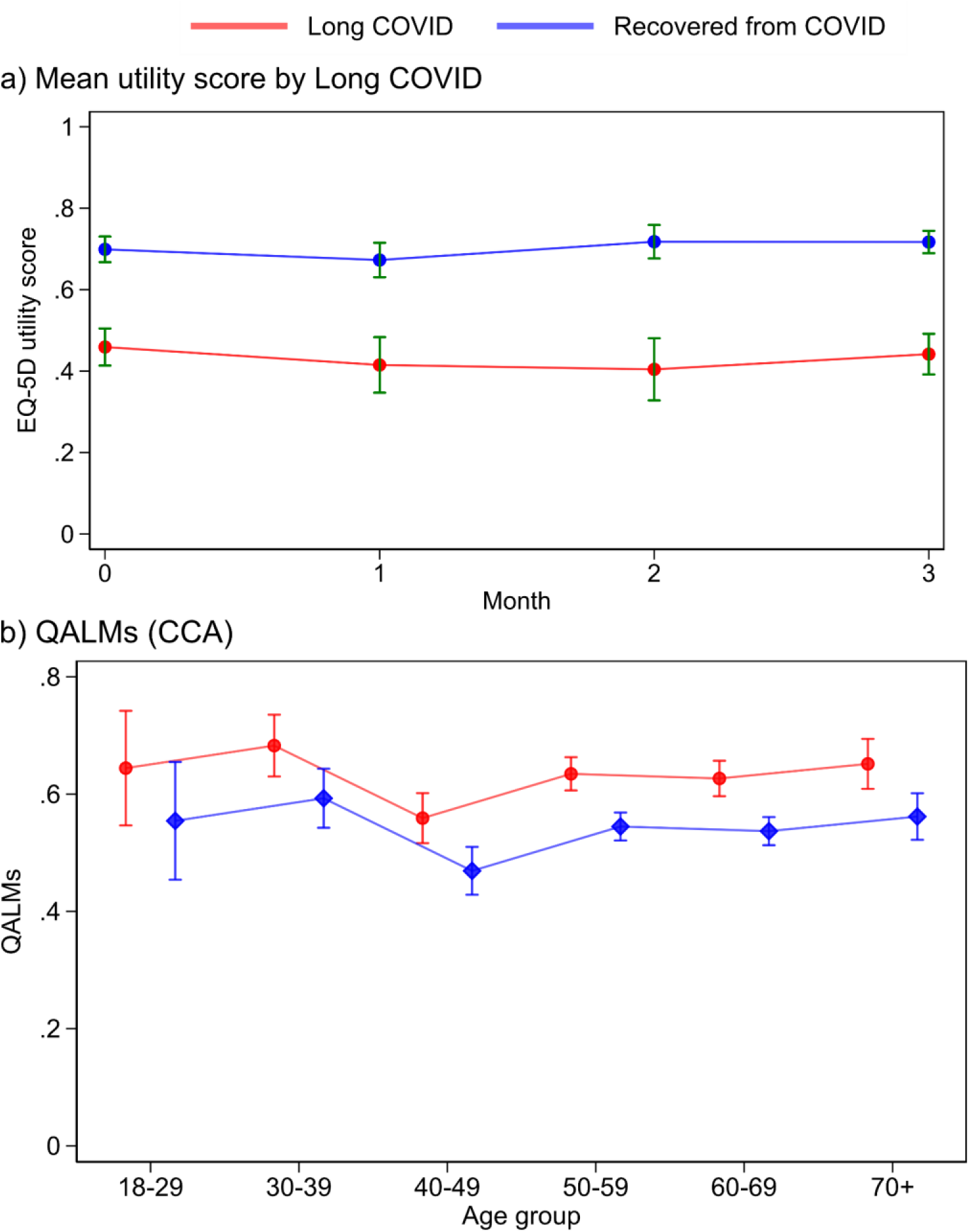
a) mean utility score in the long COVID vs non-long-COVID groups. Error bars mark 95% confidence intervals. b) Predicted Quality-adjusted Life-Months stratified by long COVID status in the complete case analysis (CCA). The linear regression model also includes a disability, number of comorbidities, and baseline utility.

We estimated QALMs using a linear regression model accounting for age, comorbidities, disability, baseline utility and sex, the predicted QALMs (Figure 4b). Lower ages represented the higher QALMs for both long COVID and recovered participants. Using the complete case approach, total QALMs for long COVID amounted to 0.85 compared to 0.44 for individuals who did not report long COVID. For available case data, QALMs amounted to 0.22 and 0.12 QALMs for long COVID and recovered participants respectively. At month 1, QALMs for long COVID respondents were between 0.253-0.271 and between months 2 and 3 this was between 0.199-0.284 (Table 2).

**Table 2:** Mean and standard deviations of quality-adjusted life-months (QALMs) stratified by long COVID for available case analysis, EHR-coded long COVID diagnosis, and complete case analysis (CCA). sd is standard deviation.

|  | Available case |  | Long COVID Diagnosis |  | Complete case |  |
| --- | --- | --- | --- | --- | --- | --- |
|  | Long COVID | No long COVID | Long COVID | No long COVID | Long COVID | No long COVID |
|  | Mean (sd) |  |  |  |  |  |
| **1 Month** | 0.25<br>(0.15) | 0.13<br>(0.12) | 0.26<br>(0.14) | 0.17<br>(0.15) | 0.26<br>(0.13) | 0.16<br>(0.13) |
| **2 Months** | 0.2<br>(0.13) | 0.11<br>(0.11) | 0.17<br>(0.12) | 0.13<br>(0.13) | 0.28<br>(0.14) | 0.15<br>(0.12) |
| **3 Months** | 0.19<br>(0.14) | 0.10<br>(0.09) | 0.21<br>(0.12) | 0.12<br>(0.12) | 0.27<br>(0.16) | 0.13<br>(0.12) |
| **Total** | 0.22<br>(0.31) | 0.12<br>(0.20) | 0.22<br>(0.3) | 0.12<br>(0.20) | 0.81<br>(0.41) | 0.44<br>(0.36) |

## Discussion

We have shown the impact of self-reported long COVID on HRQoL using a novel cohort study, which linked PROMs directly to the patient’s EHR. The mean EQ-5D score for those who self-reported long COVID was 0.49 compared to 0.71 among those without long COVID. The difference exceeds the 0.063 minimally important difference expected for EQ-5D-5L populations (McClure et al., 2017). Reported disabilities and number of diagnosed comorbidities were also associated with lower quality-of-life. The burden of long COVID was greatest in the working-age population, with higher QALMs for respondents who will participate in the labour market for longer (18-40). Comparing the utility of the whole cohort to the population norms estimated in McNamara et al. (2023), quality-of-life scores were slightly lower for non-long COVID individuals in OpenPROMPT than the population norm score of between 0.798-0.791 for participants at the same age.

Sociodemographic groups of similar characteristics to our cohort appear more likely to respond to research related to long COVID (Thompson et al., 2022; Holt et al., 2022). We found that when controlling for long COVID, participants at higher levels of socioeconomic status reported a substantially lower impact on HRQoL. Research on non-pandemic health-related conditions has found consistent inequalities across socioeconomic status, incurring higher associated medical costs, lower life expectancy and higher mortality in people of lower socioeconomic status (Maheswaran et al., 2015). To prevent similar relationships developing for long COVID, public health interventions should attempt to address the inequalities in order to prevent the gap widening following a global pandemic.

Together, breathlessness and fatigue appear to be major contributors for the decreased HRQoL attributable to long COVID, evidenced by the substantial reduction in odds ratios when FACIT-F and MRC Dyspnoea scales are included. This indicates that EQ-5D may be unable to capture the impact within the stated dimensions of quality-of-life in a population heavily impacted by both symptoms. Our results compare similarly to previous use of FACIT-F in a population of post-COVID-19 syndrome (PCS) patients, with a mean non-reversed F-score of 20.67 (SD: 12.12) slightly better than 19.6 in Walker et al., (2023), both significantly lower than the population norm value of 43.5 (Montan et al., 2018). Given FACIT-F scores were found to be highly significant on HRQoL, as suggested within Sandler et al. (2021), further assessment is needed in the interpretation of fatigue in post-COVID-19 syndrome when using EQ-5D measurements.

The EQ-5D quality-of-life index scores at baseline for self-reported long COVID participants in OpenPROMPT (0.49, SD: 0.31) were lower compared to some previous long COVID research. This is consistent across studies who have followed up patients referred to post-COVID syndrome clinics in the UK (mean 0.54, SD 0.26) (Walker et al., 2023), online surveys completed in Belgium by self-reported PCS patients (mean 0.57, SD 0.23) (Moens et al., 2022), and in previously hospitalised patients in Iran (mean 0.61, SD 0.006) (Arab-Zozani et al., 2020). EQ-5D index scores are similar to a study of patients defined as very severely impacted in physical and mental impairment by combining responses to symptom questionnaires and physical performance tests approximately 6 months after COVID-19 hospitalisation in the UK (mean 0.43, SD 0.27) which also highlighted the impact of a disability (Evans et al., 2021). Our results are therefore striking, supporting evidence from PPIE sessions where a subset of participants reported experiencing limited HRQoL over multiple years with little recovery.

To put in perspective the loss of HRQoL from long COVID, our results showed lower EQ-5D scores than from patients experiencing heart failure (mean 0.60) (Squire et al., 2017), multiple sclerosis (mean 0.59) (Carney et al., 2018), and end-stage renal disease (mean 0.68) (Yang et al., 2015).

### Strengths and limitations

A key strength of this study was the linkage of PROMS with the EHR in OpenSAFELY. This allowed more granular research using the EHR with variables such as income (which is not routinely available in EHRs), reduced the burden on participants of collecting extensive information about their medical history, and has enabled validation of collected data against the EHR, for example the number of COVID infections and hospitalisations. Combining patient responses in a trusted research environment directly into EHRs is a novel research method, with flexible tooling of questionnaires helping provide both additional information and demonstrating the convergence with medical histories. Our previous research demonstrated the difficulty of utilising only EHR data in long COVID research (Walker et al., 2021; Henderson et al., 2023), and the present study shows further limitations of EHR data for this condition: fewer than 10% of the cohort had a recorded diagnosis of long COVID in their EHR, compared to roughly 25% self-reporting long COVID.

Our cohort of 6070 respondents included in the analysis is larger than previous research on HRQoL in long COVID populations, with a higher proportion of self-reported long COVID respondents (Walker et al., 2023; Heightman et al. 2021). By collecting information on HRQoL using the validated EuroQoL EQ-5D questionnaire, and using validated instruments on breathlessness and fatigue, we were able to determine the contribution of these symptoms to the impact on quality-of-life. However, other symptoms of long COVID (or their severity) were not included in the study. Given the difficulties associated with measuring fatigue in EQ-5D-5L, it is possible that other symptoms are not well suited to measurement across the five dimensions of HRQoL in a population affected by post-COVID-19 syndrome.

Advertising for the study was among the general population and long COVID groups. Importantly, the cohort was self-selected and therefore people with long COVID, or more severe long COVID, may have been more likely to participate. Conversely, those with the most severe long COVID may have been unable to participate due to their symptoms. This study faces a similar weakness to other recruited study cohorts investigating long COVID, whereby individuals of low socioeconomic deprivation are more likely to participate. Given our and other evidence (Shabnam et al., 2023) there is a socioeconomic relationship with the risk of long COVID, the demographics of the cohort do not show this in the sample population. These factors may introduce selection bias and impact the generalisability of the findings to all long COVID patients.

The cohort also experienced high loss to follow-up and it is possible that recovery from COVID or long COVID led to loss of interest in completion across the full 90 days. This may mean that any over-representation of long COVID participants in the cohort was exaggerated over time. The lack of a reduction in QALMs lost due to long COVID over time may be attributable to this over-representation, or could have been that the data collection period was too short: for patients suffering long-term symptoms, 3 months is a relatively short period where recovery would be unexpected (Ballouz et al., 2023). Conversely, due to the symptoms of long COVID, loss to follow-up may have been due to fatigue, driven by severe long COVID symptoms. Further development of QALYs lost attributable to long COVID should consider the framework set out within Martin et al., (2021) that separates populations into clearly defined subgroups of long COVID vs. acute COVID-19 when long-term measurements of HRQoL become available. This relies on the level of missingness decreasing as long-term assessments of long COVID become more common.

A further limitation is our definition of long COVID, which was based on questions relating to recovery from long COVID and the most recent COVID episode. In order to piece together HRQoL trajectories of long COVID, it would also have been useful to know the date of the episode of COVID which led to long COVID, if the participant is able to determine this.

### Implications

Long COVID has a major impact on HRQoL in our cohort, comparatively worse than in patients experiencing heart failure (Squire et al., 2017). The effect was attenuated after adjusting for breathlessness and fatigue, indicating that these symptoms are partly responsible for the impact of long COVID on quality-of-life. This resonates with the input from our PPIE activity.

Given that the burden of long COVID on HRQoL was heaviest in the working age population, our results indicate important implications for health services, due to higher healthcare utilisation, and the wider economy. Greater economic costs can be incurred for a substantial part of the UK workforce, with some individuals reducing their labour output, and others leaving the labour market altogether (Reuschke & Houston, 2022).

Importantly, we know that there is a subset of people with long COVID that experience debilitating symptoms, and our study had participants with long COVID whose HRQoL was in a state ‘worse than death’. Though these represent a minority of people with long COVID, it is vital that support is provided to these people.

### Conclusions

Self-reported long COVID had a significant effect on quality-of-life across models accounting for different demographics. Consistent low HRQoL scores being reported across the 3-months by participants with long COVID indicates the need for targeted interventions for a cohort experiencing symptoms upwards of a year. Fatigue and the relationship with EQ-5D requires further specific research on the effectiveness of validated PROMs to capture the severity of a significant symptom in a developing condition with little current clinical treatment.

## Administrative

### Patient and Public Involvement

This study had patient and public involvement from an advisory panel of three individuals with different experiences of long COVID which we met with every 6 months. To obtain feedback on the study, separate PPI events were held in January and September 2023. LSHTM developed a website with information about OpenPROMPT, how to take part and how to contact us regarding the project (LSHTM, 2022). OpenSAFELY have developed a publicly available website (https://opensafely.org/) through which we invite any patient or member of the public to contact us regarding this study or the broader OpenSAFELY project.

## Data Availability

LSHTM is the data controller of OpenPROMPT data. NHS England is the data controller of the NHS England OpenSAFELY COVID-19 Service. TPP is the data processor; all study authors using OpenSAFELY have the approval of NHS England. This implementation of OpenSAFELY is hosted within the TPP environment which is accredited to the ISO 27001 information security standard and is NHS IG Toolkit compliant.
Access to the underlying identifiable and potentially re-identifiable pseudonymised electronic health record data is tightly governed by various legislative and regulatory frameworks, and restricted by best practice. The data in the NHS England OpenSAFELY COVID-19 service is drawn from General Practice data across England where TPP is the data processor.

## Acknowledgements

We are grateful for support from the TPP Technical Operations team throughout this work, and for generous assistance from the information governance and database teams at NHS England and the NHS England Transformation Directorate.

## Information governance and ethical approval

This research is part of the OpenPROMPT study “Quality-of-life in patients with long COVID: harnessing the scale of big data to quantify the health and economic costs” which has ethical approval from HRA and Health and Care Research Wales (HCRW) (IRAS project ID 304354). The Study Coordination Centre has obtained approval from the LSHTM Research Ethics Committee (ref 28030), as well as a favourable opinion from the South Central—Berkshire B Research Ethics Committee (ref 22/SC/0198).

LSHTM is the data controller of OpenPROMPT data. NHS England is the data controller of the NHS England OpenSAFELY COVID-19 Service. TPP is the data processor; all study authors using OpenSAFELY have the approval of NHS England (NHS Digital, 2023a). This implementation of OpenSAFELY is hosted within the TPP environment which is accredited to the ISO 27001 information security standard and is NHS IG Toolkit compliant (NHS Digital, 2023b)

Patient data has been pseudonymised for analysis and linkage using industry standard cryptographic hashing techniques; all pseudonymised datasets transmitted for linkage onto OpenSAFELY are encrypted; access to the NHS England OpenSAFELY COVID-19 service is via a virtual private network (VPN) connection; the researchers hold contracts with NHS England and only access the platform to initiate database queries and statistical models; all database activity is logged; only aggregate statistical outputs leave the platform environment following best practice for anonymisation of results such as statistical disclosure control for low cell counts (NHS Digital, 2022a).

The service adheres to the obligations of the UK General Data Protection Regulation (UK GDPR) and the Data Protection Act 2018. The service previously operated under notices initially issued in February 2020 by the Secretary of State under Regulation 3(4) of the Health Service (Control of Patient Information) Regulations 2002 (COPI Regulations), which required organisations to process confidential patient information for COVID-19 purposes; this set aside the requirement for patient consent (GOV.UK, 2022). As of 1 July 2023, the Secretary of State has requested that NHS England continue to operate the Service under the COVID-19 Directions 2020 (NHS Digital, 2022b). In some cases of data sharing, the common law duty of confidence is met using, for example, patient consent or support from the Health Research Authority Confidentiality Advisory Group (NHS HRA, n. d).

Taken together, these provide the legal bases to link patient datasets using the service. GP practices, which provide access to the primary care data, are required to share relevant health information to support the public health response to the pandemic, and have been informed of how the service operates.

## Data access and verification

Access to the underlying identifiable and potentially re-identifiable pseudonymised electronic health record data is tightly governed by various legislative and regulatory frameworks, and restricted by best practice. The data in the NHS England OpenSAFELY COVID-19 service is drawn from General Practice data across England where TPP is the data processor.

TPP developers initiate an automated process to create pseudonymised records in the core OpenSAFELY database, which are copies of key structured data tables in the identifiable records. These pseudonymised records are linked onto key external data resources that have also been pseudonymised via SHA-512 one-way hashing of NHS numbers using a shared salt. University of Oxford, Bennett Institute for Applied Data Science developers and PIs, who hold contracts with NHS England, have access to the OpenSAFELY pseudonymised data tables to develop the OpenSAFELY tools.

These tools in turn enable researchers with OpenSAFELY data access agreements to write and execute code for data management and data analysis without direct access to the underlying raw pseudonymised patient data, and to review the outputs of this code. All code for the full data management pipeline — from raw data to completed results for this analysis — and for the OpenSAFELY platform as a whole is available for review at github.com/OpenSAFELY.

The data management and analysis code for this paper was led by OC and contributed to by ADH.

