## Supplementary Appendix for "The impact of Long COVID on Health-Related Quality-of-Life using OpenPROMPT"

### Appendix: The Impact of Long COVID on HRQoL using OpenPROMPT

#### Supplementary methods

##### OpenPROMPT

OpenPROMPT was supported by the National Institute for Health and Care Research (NIHR) as a collaboration between the London School of Hygiene and Tropical Medicine, the OpenSAFELY data platform at the University of Oxford, and The Phoenix Partnership (TPP), which supplies software to General Practices in the UK. Any adult in England could participate in the study, provided they were able to download and use the Airmid smartphone application. Participants could join either passively, discovering the study page in the Airmid Research Module, or through active discovery from Airmid push notifications which invited them towards the OpenPROMPT study page. General Practices using TPP SystmOne software were invited to participate in disseminating the study to patients in their system. The study was also shared on social media to promote both the general population and individuals highly impacted by long COVID to enrol.

##### Long COVID Histories in Electronic Health Records (EHRs)

Alongside the questionnaire data collection, histories related to COVID-19 in medical records utilised codes created in OpenSAFELY. For the comparison between the numbers self-reporting long COVID to diagnosed cases in EHRs, definition of a long COVID diagnosis was based on SNOMED CT codes from NICE guidelines on managing the long-term effects of COVID-19:

| SNOMED CT Code | |
|---|---|
| 1325161000000102 | Post-COVID-19 syndrome |
| 1325181000000106 | Ongoing symptomatic disease caused by severe acute respiratory syndrome coronavirus 2 |

For wider assessment of long COVID histories, the assessment of any record relating to long COVID was based on codes including referral to specialist long COVID clinics:

| SNOMED CT Code | |
|---|---|
| 1325021000000106 | Signposting to Your COVID Recovery |
| 1325031000000108 | Referral to post-COVID assessment clinic |
| 1325041000000104 | Referral to Your COVID Recovery rehabilitation platform |
| 1325051000000101 | Newcastle post-COVID syndrome Follow-up Screening Questionnaire |
| 1325061000000103 | Assessment using Newcastle post-COVID syndrome Follow-up Screening Questionnaire |
| 1325071000000105 | COVID-19 Yorkshire Rehabilitation Screening tool |

| | |
|---|---|
| 1325081000000107 | Assessment using COVID-19 Yorkshire Rehabilitation Screening tool |
| 1325091000000109 | Post-COVID-19 Functional Status Scale patient self-report |
| 1325101000000101 | Assessment using Post-COVID-19 Functional Status Scale patient self-report |
| 1325121000000105 | Post-COVID-19 Functional Status Scale patient self-report final scale grade |
| 1325131000000107 | Post-COVID-19 Functional Status Scale structured interview final scale grade |
| 1325141000000103 | Assessment using Post-COVID-19 Functional Status Scale structured interview |
| 1325151000000100 | Post-COVID-19 Functional Status Scale structured interview |

Hospitalisation with COVID-19 was defined using data from Hospital Episode Statistics Admitted Patient Care (HES-APC) accessed through OpenSAFELY. The dataset used ICD-10 codes to define any previous COVID-19 related hospitalisation.Though U072 states that COVID-19 was not identified, the code was used when suspected but testing proved inconclusive:

| icd10_code | |
|---|---|
| U071 | covid19 virus identified |
| U072 | covid19 virus not identified |

#### Supplementary figures

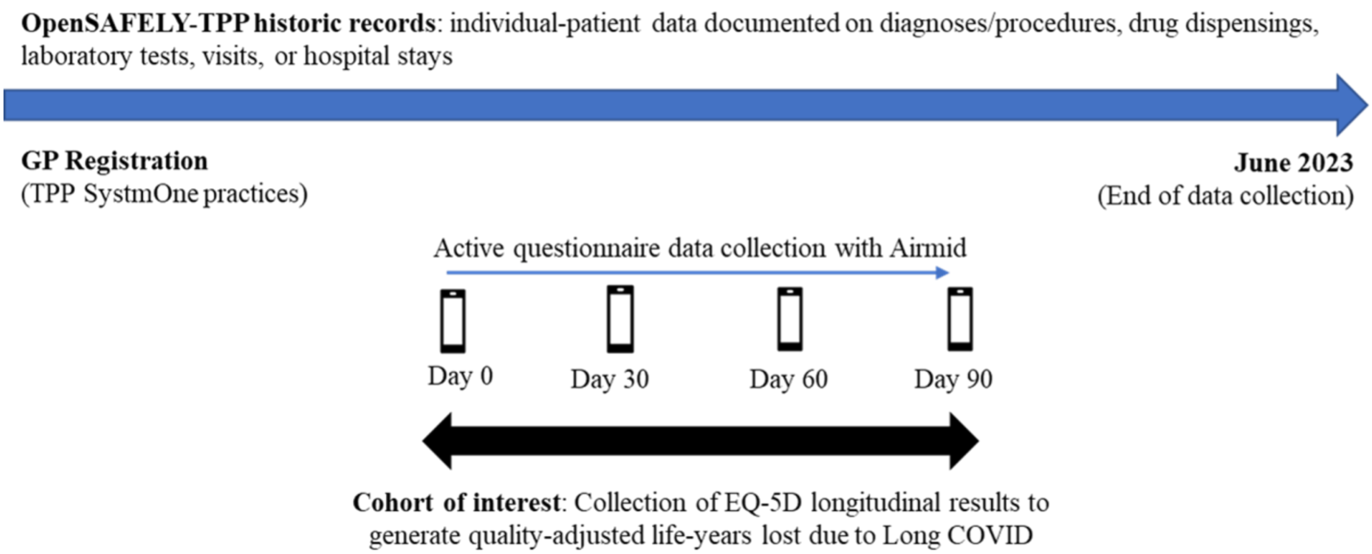

**Supplementary figure 1.** Study design diagram of the primary cohort of OpenPROMPT.

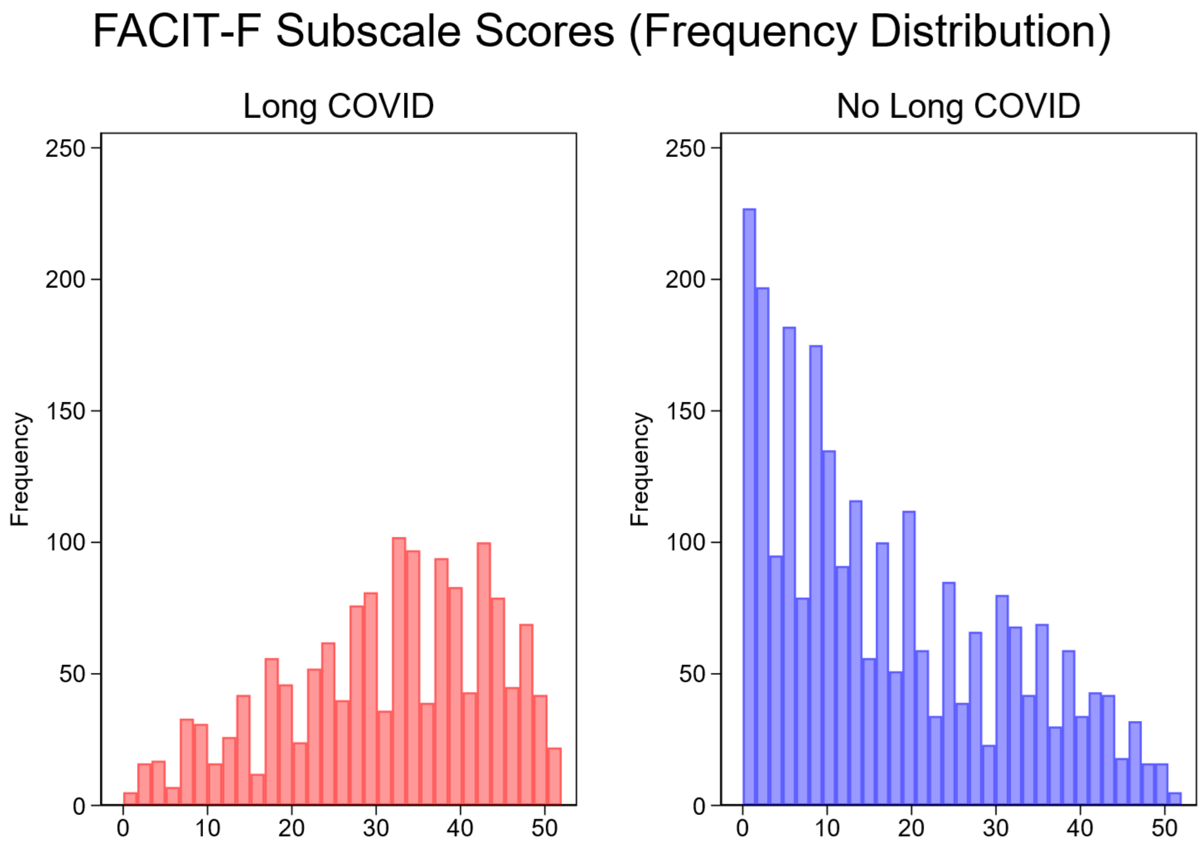

**Supplementary figure 2.** Frequency distribution of reversed FACIT-F fatigue scores at recruitment survey stratified by self-reported long COVID. Higher scores indicate greater fatigue

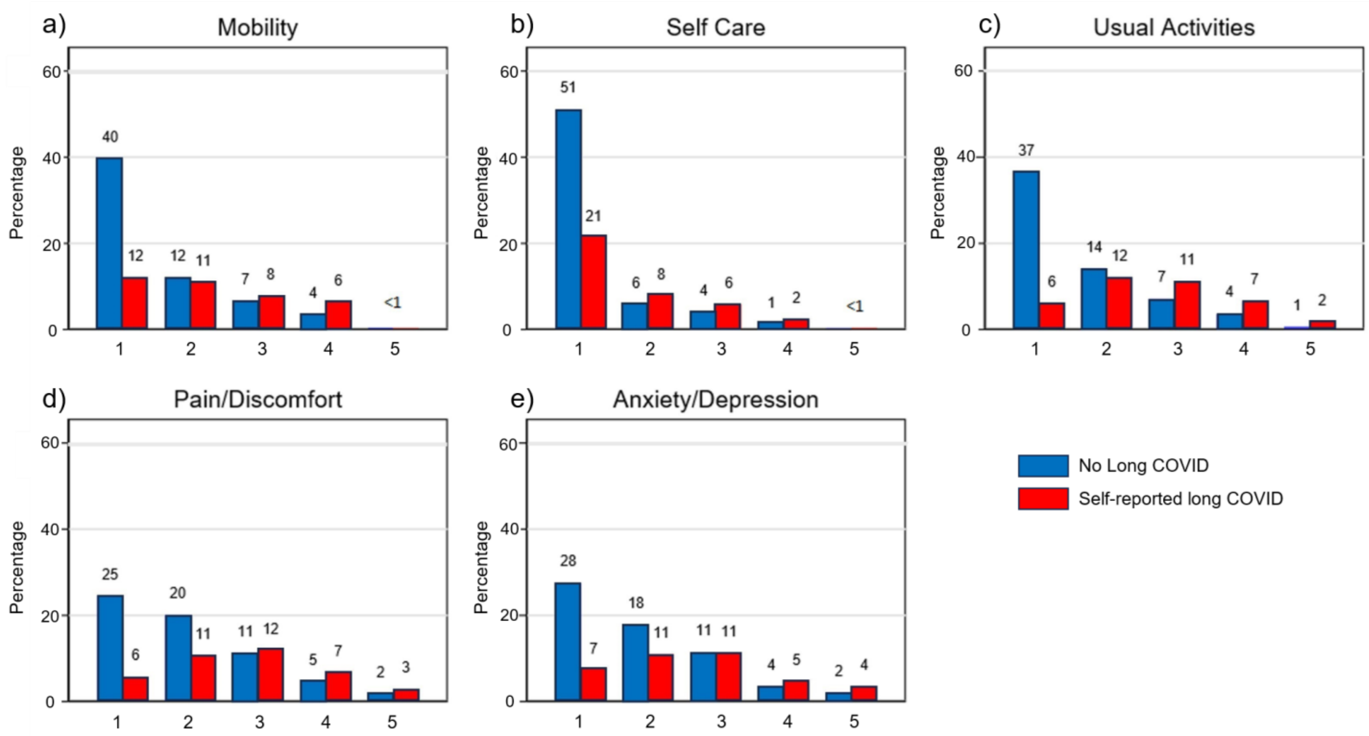

**Supplementary figure 3.** Self-reported quality-of-life measures. Percentage responses for the five dimensions of HRQoL measured in EQ-5D-5L. a) Mobility dimension of ED-5D, b) Self-care dimension of EQ-5D, c) Usual activities dimension of ED-5D. d) Pain/discomfort dimension of ED-5D, e) Anxiety/depression dimension of ED-5D. Each dimension has five possible responses: level 1: no problems, level 2: slight problems, level 3: moderate problems, level 4: severe problems, and level 5: extreme problems/unable to. Blue marks the participant did not report long COVID, and red that they did. Responses are only shown for the 3975 non-missing self-reported long COVID respondents

#### Supplementary tables

**Supplementary table 1.** List of Comorbidities

| **Disease** |
| --- |
| Non-haematological cancer |
| Haematological cancer[1] |
| Chronic respiratory disease |
| Chronic cardiac disease |
| Chronic liver disease |
| Stroke or dementia |
| Other neurological conditions[2] |
| Mental health conditions[3] |
| Organ transplant |
| Rheumatoid arthritis |
| Systemic lupus erythematosus |
| Psoriasis |
| Other immunosuppressive conditions[4] |

1. Having haematological cancers six months before the index date; 2. Such as Huntington's disease, multiple sclerosis, motor neuron diseases, and other neurological diseases; 3. Consists of psychosis, schizophrenia, bipolar disorder, and depression; 4. Including other permanent and temporary immunosuppressive diseases.

| | First Part<br>Probability of HRQoL loss | | Second Part<br>Unit loss in HRQoL | |
|---|---|---|---|---|
| | Odds ratio | 95% CI | Coefficients | 95% CI |
| **Self-Reported long COVID** | 21.2*** | [12.03, 37.54] | 0.075*** | [0.06, 0.09] |
| **Males** | 0.9 | [0.61, 1.31] | -0.021* | [-0.04, 0.00] |
| **Age (base 18-29)** | | | | |
| 30-39 | 0.39 | [0.14, 1.08] | 0.027 | [0.00, 0.06] |
| 40-49 | 0.31* | [0.12, 0.81] | 0.046** | [0.02, 0.08] |
| 50-59 | 0.19*** | [0.07, 0.48] | 0.039* | [0.01, 0.07] |
| 60-69 | 0.12*** | [0.05, 0.33] | -0.023 | [-0.05, 0.01] |
| 70+ | 0.14*** | [0.05, 0.42] | -0.023 | [-0.06, 0.02] |
| **Number of Comorbidities (Base 0)** | | | | |
| 1 | 2.18*** | [1.45, 3.29] | 0.023** | [0.01, 0.04] |
| 2 | 2.85* | [1.25, 6.49] | 0.042** | [0.01, 0.07] |
| 3+ | 0.90 | [0.16, 5.14] | 0.10*** | [0.05, 0.16] |
| **Disabled** | 60.2*** | [27.79, 130.57] | 0.28*** | [0.26, 0.30] |
| **Highest Education (Base College/University)** | | | | |
| Primary School/Less | 1 | [1.00, 1.00] | 0.051 | [-0.04, 0.15] |
| Secondary/high school | 1.05 | [0.66, 1.68] | 0.029** | [0.01, 0.05] |
| Postgraduate qualification | 1.14 | [0.74, 1.76] | -0.026** | [-0.04, -0.01] |
| **Household Income (Base £32000-47999)** | | | | |
| £6000-12999 | 5.84*** | [2.1, 16.27] | 0.12*** | [0.09, 0.15] |
| £13000-18999 | 1.47 | [0.67, 3.24] | 0.095*** | [0.07, 0.12] |
| £19000-25999 | 1.26 | [0.66, 2.40] | 0.038** | [0.01, 0.06] |
| £26000-31999 | 1.64 | [0.85, 3.18] | 0.027* | [0.00, 0.05] |
| £48000-63999 | 0.81 | [0.46, 1.43] | -0.011 | [-0.04, 0.01] |
| £64000-95999 | 0.87 | [0.48, 1.59] | -0.015 | [-0.04, 0.01] |
| £96000 + | 0.44* | [0.22, 0.89] | -0.017 | [-0.05, 0.02] |
| **IMD Quintiles (Base 1st (most deprived))** | | | | |
| 2nd | 0.52 | [0.25, 1.10] | 0.014 | [-0.01, 0.04] |
| 3rd | 0.51 | [0.25, 1.04] | -0.02 | [-0.04, 0.00] |
| 4th | 0.39* | [0.19, 0.81] | -0.03* | [-0.05, -0.01] |
| 5th (least deprived) | 0.40* | [0.19, 0.81] | -0.029* | [-0.05, 0.00] |
| **Constant** | 41.7*** | [12.9, 135.03] | 0.23*** | [0.19, 0.26] |

**Supplementary table 2.** Multivariable Mixed effect regression models on the probability of HRQoL loss, and the resulting unit loss associated with covariates (Note: negative coefficients indicate better HRQoL). The log of variance was 7.77*** (95% CI: 5.52-10.94). The number of observations was 5253 for the first part, and 4483 for the second part. Akaike Information Criterion (AIC) for the first part was 3240.9 and -2452.3 for the second part. * p<0.05, ** p<0.01, *** p<0.001

Disabled participants had the greatest probability of reporting an impact on quality-of-life (OR for returning a loss of HRQoL: 60.2 (95% CI: 27.79,130.57), and an estimated 0.28 (CI: 0.26; 0.30) unit lower quality-of-life (supplementary table 2). Those with comorbidities reported an impact on quality-of-life, especially evident for one comorbidity (OR:2.18, CI:1.45, 3.29) and decreasing as comorbidities increase. The unit decrease in quality-of-life was significant for all levels in the second part, between 0.023-0.1. No effect was found on the impact of ethnicity across models. Evidence was found on the probability of lower ages reporting worsened quality-of-life**.** Lower quintiles of deprivation and higher levels of household income were found to improve quality-of-life compared to baseline levels. No evidence was found on the probability of education level predicting changes in quality-of-life, but there was a small impact of lower levels having higher unit decreases in quality-of-life.

| | First Part<br>Probability of HRQoL loss | | Second Part<br>Unit loss of HRQoL | |
|---|---|---|---|---|
| | **Odds ratio** | **95% CI** | **Coefficients** | **95% CI** |
| **Self reported long COVID** | 2.35*** | [1.37, 3.61] | 0.00093 | [-0.01, 0.01] |
| **Disabled** | 7.49*** | [4.04, 13.89] | 0.16*** | [0.15, 0.17] |
| **Male** | 1.3 | [0.93, 1.83] | 0.0027 | [-0.01, 0.01] |
| **Age (Base 18-29)** | | | | |
| 30-39 | 0.26** | [0.11, 0.65] | 0.018 | [0.00, 0.04] |
| 40-49 | 0.31** | [0.13, 0.71] | 0.028** | [0.01, 0.05] |
| 50-59 | 0.31** | [0.14, 0.70] | 0.038*** | [0.02, 0.06] |
| 60-69 | 0.35* | [0.15, 0.80] | 0.030** | [0.01, 0.05] |
| 70+ | 0.37* | [0.15, 0.92] | 0.029* | [0.00, 0.06] |
| **Number of Comorbidities (Base 0)** | | | | |
| 1 | 1.45* | [1.01, 2.08] | 0.0067 | [0.00, 0.02] |
| 2 | 1.32 | [0.65, 2.71] | 0.0044 | [-0.02, 0.02] |
| 3+ | 0.23 | [0.04, 1.26] | 0.005 | [-0.03, 0.04] |
| **MRC Breathlessness (Base grade 1)** | | | | |
| Grade 2 | 2.89*** | [2.04, 4.08] | -0.0077 | [-0.02, 0.00] |
| Grade 3 | 5.27*** | [2.22, 12.48] | 0.037*** | [0.02, 0.05] |
| Grade 4 | 5.65 | [0.57, 56.40] | 0.13*** | [0.11, 0.15] |
| Grade 5 | 1 | [1.00, 1.00] | 0.26*** | [0.23, 0.29] |
| **FACIT-F score** | 1.26*** | [1.22, 1.30] | 0.0094*** | [0.01, 0.01] |
| **Constant** | 0.82 | [0.38, 1.79] | 0.066*** | [0.04, 0.09] |

**Supplementary table 3.** Multivariable Mixed effect regression models on the probability of HRQoL loss with the inclusion of symptom specific patient-reported outcome measures (PROMs), and the resulting unit loss associated with covariates (Note: negative coefficients indicate better HRQoL). The log of variance was 5.20*** (95% CI: 3.52-7.69). The number of observations was 6181 for the first part, and 5433 for the second part. Akaike Information Criterion (AIC) for the first part was 3112.8 and -4850.7 for the second part. * p<0.05, ** p<0.01, *** p<0.001

| | QALYs | |
|---|---|---|
| | Long COVID | No Long COVID |
| | Mean (SD) | |
| **1 Month** | 0.043 (0.022) | 0.027 (0.022) |
| **2 Months** | 0.047 (0.024) | 0.025 (0.02) |
| **3 Months** | 0.046 (0.026) | 0.022 (0.02) |
| **Total** | 0.135 (0.068) | 0.073 (0.061) |

**Supplementary table 4.** Individual level Quality-Adjusted life-years (QALYs) stratified by long COVID for complete cases, calculated using Area Under the Curve (AUC). These are reshaped from the Quality-Adjusted life-months (QALMs) to reflect life-years.
